# Olfactory training using nasal inserts more effective due to increased adherence

**DOI:** 10.1101/2024.06.26.24309521

**Authors:** Anja L. Winter, Sofie Henecke, Evelina Thunell, Mattias Swartz, Joakim Martinsen, Pernilla Sahlstrand Johnson, Johan N. Lundström

**Affiliations:** Department of Clinical Neuroscience, Karolinska Institutet, Stockholm, Sweden; Skåne University Hospital, Department of Oto-Rhino-Laryngology, Malmö, Sweden; Lund University, Department of Clinical Sciences, Malmö, Sweden; Department of Otorhinolaryngology, Karolinska University Hospital, Stockholm, Sweden; Monell Chemical Senses Center, Philadelphia, PA, United States

## Abstract

The recommended treatment for hyposmia (clinically reduced sense of smell) is olfactory training using odors in containers that the patients smell twice a day for several weeks. Adherence to the olfactory training regimen is, however, generally low. We aimed to investigate if a new form of olfactory delivery via scented nasal inserts could enhance olfactory training adherence by allowing participants to be mobile while performing the training and thereby lower perceived intrusion in their everyday life, using a randomized controlled parallel-group design. Two groups of individuals with hyposmia underwent 8 weeks of olfactory training. One group (n=60) performed olfactory training using scented nasal inserts (nasal plugs that retain nasal patency) and one group (n=56) performed the standard care regiment currently recommended by the Swedish healthcare system. We assessed objective and subjective olfactory ability before and after olfactory training as well as adherence to training. While both groups significantly improved their objective and subjective olfactory abilities, training with nasal insert produced similar improvement as standard care in overall treatment outcome. However, there was a significantly greater increase in discrimination performance and lower dropout rate (6.7%) in the nasal insert compared to the standard care group (23.2%). Critically, the nasal insert group had a significantly higher adherence to the training regimen, i.e. fewer missed training sessions. In addition, they reported overall greater satisfaction with their treatment. These data suggest that olfactory training with nasal inserts could serve as a more effective form of treatment for hyposmia, due to patients improved adherence to protocol and increased tendency to finish their treatment regimen.

## INTRODUCTION

Olfactory training, sometimes also referred to as smell training, involves repetitive, regular, and time extended purposeful exposure to odors and is currently the most commonly recommended treatment regimen for patients with olfactory dysfunction (Whitcroft et al., 2023). Patients are typically instructed to smell a number of either provided odors or common household odors for 15 to 20 minutes, twice a day, for one to three months, or longer (Hummel et al., 2009). This cost-effective treatment has been considered fairly efficient, with a majority of patients who complete their treatment experiencing improved olfactory function (Huang et al., 2021; Hummel et al., 2009; Kattar et al., 2021; Sorokowska et al., 2017). However, as with many forms of extended at-home treatment regimens, adherence to olfactory training is low with study dropout rates as high as 45% (Fornazieri et al., 2020; Lamira et al., 2019) and with a potentially lower compliance outside experimental studies, in everyday clinical practice.

Non-adherence to treatment is a pervasive problem for all types of at-home treatment and is observed across various healthcare disciplines. The exact extent of non-adherence to olfactory training in clinical non-experimental populations is poorly explored, but non-adherence to chronic medication has been estimated to 50% (M Robin DiMatteo et al., 2002) and non-adherence to physiotherapy, of which olfactory training could be considered one form, could be as high as 70% (Sluijs et al., 1993) in clinical non-experimental populations.

Interestingly, subjective experiences of both presence and lack of olfactory improvement during olfactory training treatment is associated with non-adherence (Haas et al., 2024). In other words, individuals stop performing the training on their own accord both if they feel that their sense of smell improves, and if they feel that it does not improve. This is problematic considering that subjective and objective olfactory performance are poorly correlated (Landis et al., 2003).

A probable reason for the high number of patients not being persistent with their treatment regime is that olfactory training, although promising in its potential to aid in the recovery of olfactory function, is often described as tedious and time-consuming and it restricts the individual to remain within one location focusing only on the task during each training session (Haas et al., 2024). Engaging in repetitive exercises while confined to one place for an extended period demands both dedication and patience, and the necessity for consistent and long-term commitment could be challenging. An easier and more manageable method of training could increase adherence and thereby also enhance treatment effects on the group level.

A potential remedy to some often-mentioned problems with olfactory training aiming to increase compliance is to enable participants to perform the training in a less intrusive manner. A recent product innovation is scented silicon nasal inserts that sit birhinally within the nostrils, combined by a small silicon bridge, yet allowing near normal nasal patency. These are currently used to mask negative external odors but could, modified to have a range of alternative odors representing distinct odor objects, potentially serve as a mobile olfactory training system.

Here, we aimed to investigate how a modified version of olfactory training using scented nasal inserts compares to standard olfactory training. The scented nasal inserts provide continuous olfactory training and liberates participants from the constraint of remaining stationary during their training sessions. This provides the freedom to engage in other everyday activities while completing the olfactory training. We hypothesized that this increased mobility would lead to increased compliance to the olfactory training regimen, and consequently better treatment outcome, as compared to standard olfactory training.

## MATERIAL AND METHODS

### Participants

Participants (*n* = 173) were recruited from two outpatient clinics and via social media advertisement. Inclusion criteria were functional hyposmia at baseline, defined as TDI between 15.25 and 31.25, and age between 18 and 65 years old. Exclusion criteria were any psychiatric diagnoses, non-viral or non-idiopathic cause of their olfactory dysfunction (such as head trauma, surgery etc.), as well as enrollment in other olfactory training studies.

After initial screening with TDI test (described below), a total of 123 participants were enrolled in the study. Seven of these were excluded before analyzes; four due to problems with the testing conditions, two due to nasal congestion, and one due to not attending the second visit. The final sample consisted of 116 individuals (Table 1) which were, at their first testing visit (baseline visit), randomized into either training with Nasal inserts (*n*=60) or Standard care (*n*=56) for eight weeks. At the baseline visit, participants reported having experienced olfactory dysfunction for an average of 41 months. All participants visited the lab on two occasions. At the baseline visit and after the 8-week long training period, their olfactory functions were assessed (see below) and they answered questionnaires on demographic information and quality of life (reported elsewhere following a future one-year follow-up). The two testing visits were seemingly identical with the exception that questions about demographics were replaced with questions regarding adherence to the training protocol. The mean duration between the first and second visit was 68 days. There were no statistical differences between groups (all *p* > .05) in age, sex, duration of olfactory dysfunction, baseline olfactory measurements scores, and time elapsed between the two visits (Table 1). All procedures were in accordance with the Helsinki declaration, approved by the Swedish Ethical Review Authority (Dnr: 2023-03779-01), and all participants provided written informed consent prior to participation.

**Table 1.** Descriptive statistics of research participants with statistical tests of difference between olfactory training groups.

| *Variable* | *Nasal Insert*<br>*(N = 60)* | *Standard Care*<br>*(N = 56)* | *Statistic* | *p* | *Total*<br>*(N = 116)* |
| --- | --- | --- | --- | --- | --- |
| Sex | | | $\chi^2 = .03$ | .86 | |
| Female (percentage) | 50 (83.3%) | 45 (80.4%) |  |  | 95 (81.9%) |
| Male (percentage) | 10 (16.7%) | 11 (19.6%) |  |  | 21 (18.1%) |
| Age | | | $t = .90$ | .37 | |
| Mean (SD) | 49.3 (12.1) | 47.5 (9.08) |  |  | 48.4 (10.7) |
| Duration hyposmia (months) | | | $t = .19$ | .85 | |
| Mean (SD) | 42.2 (41.6) | 40.6 (48.3) |  |  | 41.4 (44.7) |
| TDI visit 1 | | | $t = .16$ | .88 | |
| Mean (SD) | 24.7 (4.67) | 24.9 (4.71) |  |  | 24.8 (4.66) |
| Days between visits | | | $t = .11$ | .90 | |
| Mean (SD) | 68 (24.5) | 68.5 (24.8) |  |  | 68.2 (24.5) |

### Procedures

#### Nasal Insert Training

Participants randomized to the nasal insert olfactory training group (NI) were provided with scented nasal plugs (Nosaplugs, NosaMed AB, Stockholm; Figure 1A) that they were instructed to wear for 20 minutes in the morning and 20 minutes in the evening, every weekday for 8 weeks. Standard clinical practice recommended by the Swedish healthcare system is typically a 12-week long training regimen (Ahnblad, 2023). The nasal plugs are inserted into the bilateral nostrils (Figure 1B) and administer a specific scent (vanilla, lemon, melon, rosemary, menthol, orange, peach, strawberry, cherry, or cola) while still allowing the individual to retain near normal nasal patency. Participants used one scent for 10 minutes and then replaced it with another scent for the remaining 10 minutes of the session, resulting in a 20-minute-long odor training session, twice daily. They further visualized and focused on the smell during training which was aided by an image of the odor object printed on each individual nasal plug package. All participants were contacted twice during their 8 week at-home training period with information regarding where to reach out had they any questions, comments, or concerns: A text message was sent out one week after the initial visit, followed by a phone call approximately three weeks later.

**Figure 1.**
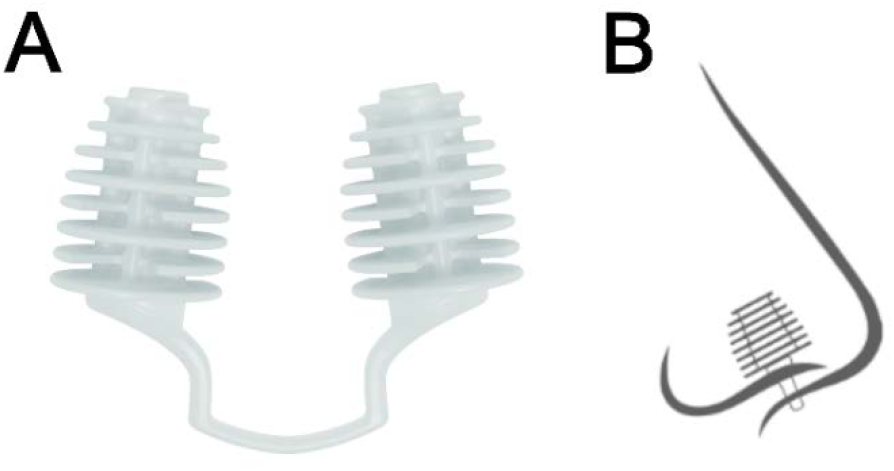
**A**. Nasal insert with its scented lamella that allow near normal nasal patency. **B**. Schematic drawing of a nasal insert positioned within the nose, viewed from the side.

#### Standard Olfactory Training

Participants in the standard care olfactory training group (SC) were instructed to choose 4-6 household odors to smell for 20 minutes in the morning and 20 minutes in the evening, every weekday for 8 weeks. This training regimen follows standard clinical practice recommended by the Swedish healthcare system with two important changes to more directly equate the training of the two groups. Instead of daily training throughout the full study period, training was only done during weekdays. Moreover, training was conducted for 8 weeks and not the recommended 12. These adjustments were done to avoid potential excessive dropout rates given that informal observations at the clinics put patient training dropout to around 80%. Participants smelled each item for 10 to 20 seconds and visualized and focused on the smell during training. Identically to the NI group, all participants were contacted twice during their 8-week at-home training period with information regarding where to reach out had they any questions, comments, or concerns. A text message was sent out one week after the initial visit, followed by a phone call approximately three weeks later. After completion of the study, participants in the SC group were offered to take home the same olfactory training kit that was provided to the NI group. Apart from the type of olfactory training, both groups received identical treatment and procedures throughout the study.

### Measurements

#### Olfactory Function

##### Objective Olfactory Function

Objective olfactory function was assessed before and after 8 weeks of olfactory training using the Sniffin’ Sticks extended test battery (Burghart Messtechnik, Holm, Germany) where pen-like tools are used to present odors and evaluate nasal chemosensory performance on odor detection threshold for phenyl ethyl alcohol (T, range 1 to 16), odor quality discrimination (D, range 1 to 16), and cued odor identification (I, range 1 to 16). Combined, these three tests generate an additive TDI score (range 3 to 48) that reflects overall olfactory function, with higher scores indicating better function.

##### Subjective Olfactory Function

Subjective olfactory function over the past three days was self-assessed using a 10-point visual analogue scale (VAS), ranging from 0 (no sense of smell) to 10 (excellent sense of smell).

### Compliance

Dropout. Dropout rate was defined per group as the percentage of individuals who did not attend the post-training visit. We obtained this number by counting the total number of participants who, at any point before their post-training visit, decided to discontinue their participation, either by informing the experimenter or ending their communication without explanation. Individuals who ended their training prematurely but still turned up at the post-training visit were not counted as dropouts.

Adherence. To measure treatment adherence, we used a five-item questionnaire (Haas et al., 2024) that participants answered during their second visit. The questionnaire separately assesses consistency, perceived tediousness, forgetfulness, and cause for potential discontinuation of the training. Specifically, the adherence questionnaire contained the following questions, “Did you consistently perform olfactory training?”, “Did you stop performing olfactory training on your own accord, because you felt that your sense of smell did not improve?”, “Did you stop performing olfactory training on your own accord, because you felt that your sense of smell did improve?”, “Did you feel that performing olfactory training twice a day was too often?”, “How often did you forget to perform olfactory training?”

### Statistical Analyses

Upon journal acceptance, all raw data and analyses scripts will be available at the Open Science Framework (OSF). The study hypothesis, inclusion/exclusion criteria and analyses plan were preregistered on clinicaltrials.gov, ID NCT06142565. Statistical analyses were performed using the statistical software R (v4.3.3; R Core Team, 2024) and the packages car (v3.1.2; Fox & Weisberg, 2019), dplyr (v1.1.4; Wickham et al., 2023), ggbeeswarm (v0.7.2; Clarke et al., 2023), ggplot (v3.5.1; Wickham, 2016), ggpubr (v0.6.0; Kassambara, 2023), haven (v2.5.4; Wickham et al., 2023), Hmisc (v5.1.2; Harrell, 2024), table1 (v1.4.3; Rich, 2023), tidyr (v1.3.1; Wickham et al., 2024), and tidyverse (v2.0.0; Wickham et al., 2019). The significance criterion for the statistical tests was set to α = 0.05.

## RESULTS

### Objective Olfactory Improvement

First, we investigated the overall effect of olfactory training on objective olfactory ability. By directly comparing the pre-test with the post-test using a dependent samples t-test, for both groups combined, we found a significant difference in TDI between the two visits, *t*(98) = 5.7, *p* < .001, demonstrating that olfactory function improved in the full sample, Figure 2. We then determined whether there was a difference in olfactory improvement between the two training groups. By using a one-way ANCOVA to compare the post-treatment TDI score between groups while using baseline TDI score as a covariate, we found no significant difference in treatment effects between the two groups after controlling for baseline TDI, *F*(1, 96) = 3.37, *p* = .07; Figure 3A. Both groups did, however, on average independently improve their olfactory performance due to training, as demonstrated by separate paired t-tests, *t*(55) = 6.33, *p* < .001 for the NI group and *t*(42) = 2.18, *p* = .035 for the SC group. On average, TDI values increased in the NI group by 3.63 (*SD* = 4.29), amounting to an average TDI increase from baseline of 15.6%, and in the SC group by 1.8 (*SD* = 5.46), an average TDI increase of 8.1%. Having established that nasal insert is a non-inferior alternative to standard care in regards of change in TDI scores, we next wanted to determine whether there was a difference in olfactory improvement between the two training groups in the three subtests. By using the same type of analysis as for the TDI, but this time for each of the three sub-tests scores, we found a significant difference in olfactory improvement for discrimination, *F*(1, 96) = 3.96, *p* = .049; Figure 3C, between the two training groups.

**Figure 2.**
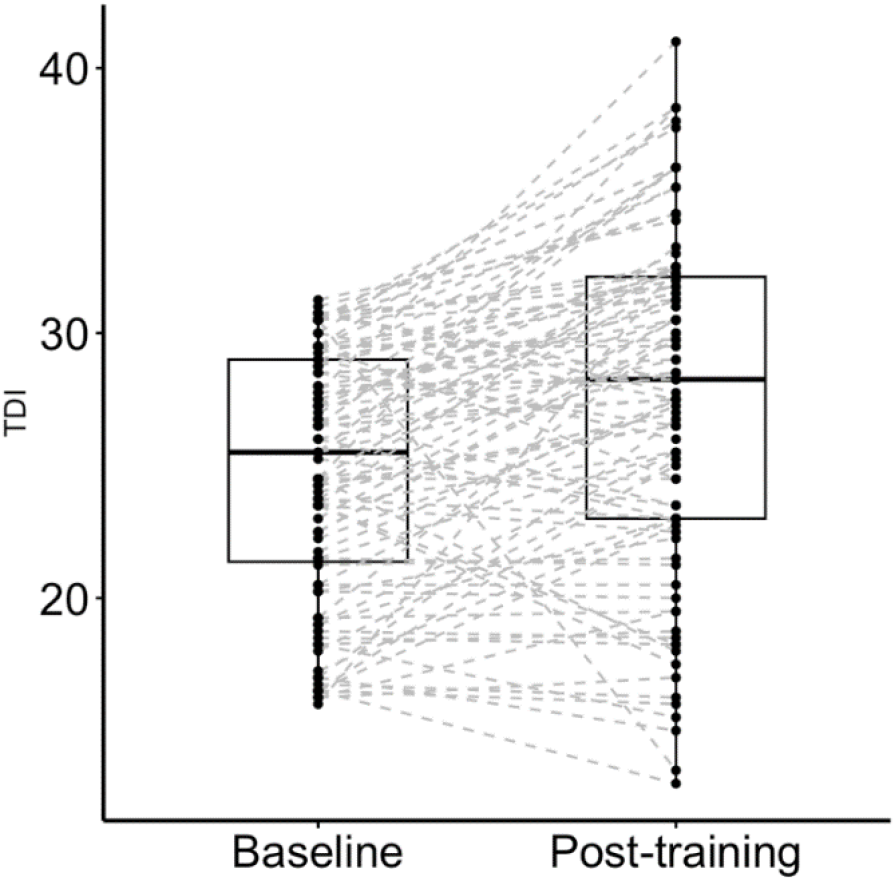
TDI scores at baseline and post-training visits with individual scores connected. Dots represent individual scores, and dashed lines connect TDI scores for the two visits for each individual. Solid bars represent group medians and boxes represent interquartile intervals

**Figure 3.**
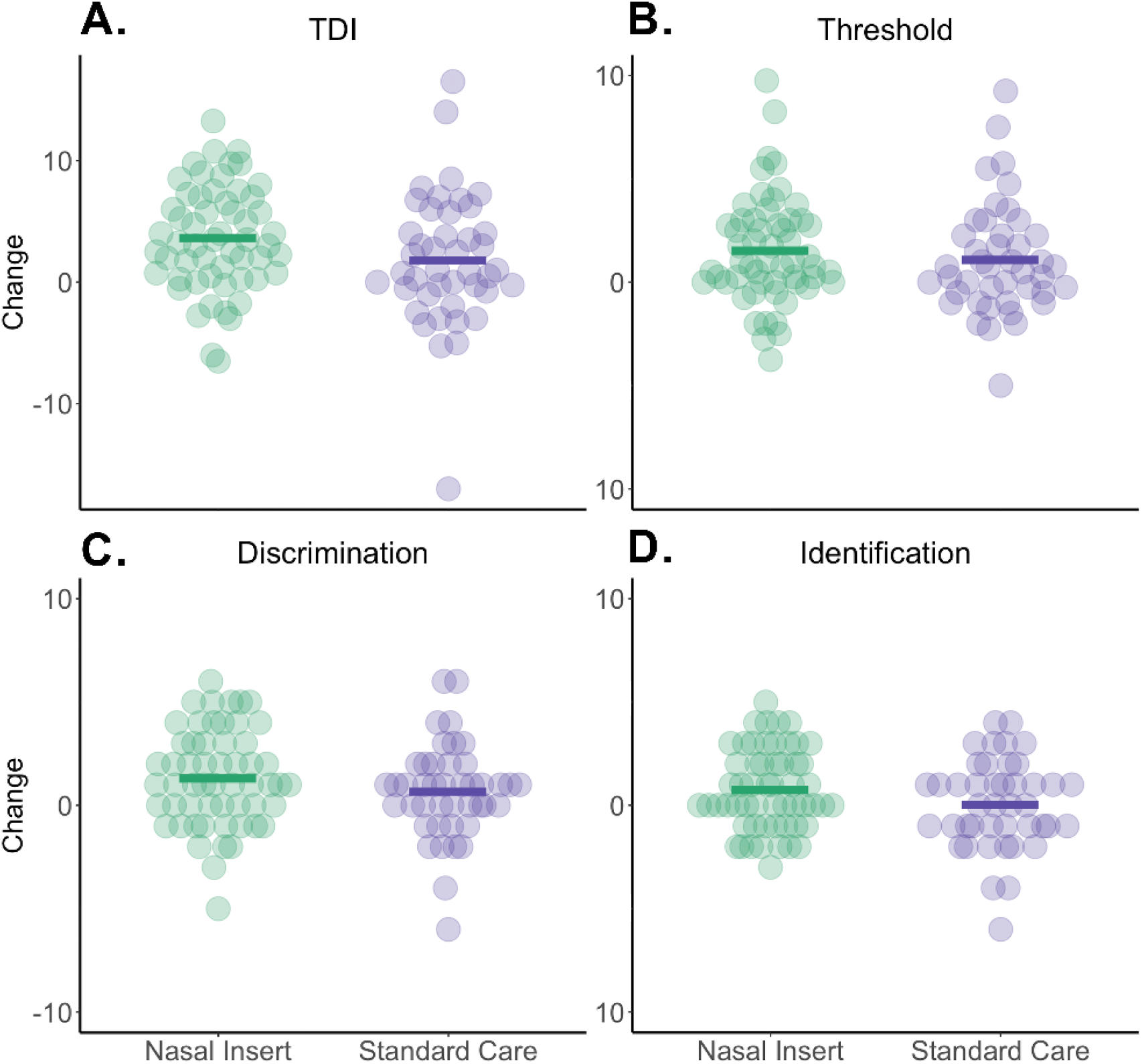
Change in objective olfactory function from baseline to post-treatment per olfactory test score and olfactory training group. **A**. Combined TDI scores. **B**. Odor detection threshold scores. **C**. Odor quality discrimination scores. **D**. Odor identification scores. In all panels, dots represent individual values and solid bars depict group means. Note the difference in scale between panels A and B-D.

However, no significant differences in olfactory improvement were detected between the two training groups for threshold, *F*(1, 96) = .7, *p* = .41; Figure 3B, or identification scores, *F*(1, 96) = 1.9, *p* = .17; Figure 3D, when controlling for each individual’s sub-test baseline score.

### Subjective Olfactory Improvement

We also wanted to estimate the overall effect of olfactory training on subjective olfactory ability. By comparing the pre-test with the post-test using a dependent samples t-test, we found a significant difference in subjective olfactory ability between the two visits, *t*(98) = 5.5, *p* < .001. We then wanted to determine whether there was a difference in subjective olfactory improvement between the two training groups. By using a one-way ANCOVA to compare the post-treatment subjective olfactory function score between groups while using baseline treatment subjective olfactory function score as a covariate, we found no significant difference in treatment effect, *F*(1, 96) = 3.5, *p* = .06.

Previous studies have shown that the duration of olfactory dysfunction has a negative impact on olfactory training treatment effect (Kattar et al., 2021). To assess this effect in our samples, we calculated the Spearman’s rank order correlation between olfactory improvement and time since onset of olfactory dysfunction in our sample. We found no significant association between the duration of dysfunction and either objective olfactory function, *r*(97) = -.12, *p* = .22, or subjective olfactory function, *r*(97) = .03, *p* = .77.

### Dropout and Treatment Adherence

We then assessed whether there were any potential differences in dropout rates and adherence to training between the two groups. Assessing dropout rates, we found that there were significantly fewer dropouts in the NI than the SC group, *X*^*2*^ (1, *N* = 99) = 5.09, *p* = .02. The total number of individuals classified as dropouts were 4 in the NI group (6.7%) and 13 in the SC group (23.2%).

Turning our focus to treatment adherence, we wanted to know whether there was a difference between the two training groups in how they responded to the adherence questionnaire. By separately comparing the answers to adherence questionnaire between the two groups using independent Chi-square tests, we found a significant difference in answers to the questions “Did you consistently perform olfactory training?”, *X*^*2*^ (1, *N* = 99) = 7.8, *p* = .005, and “Did you feel that performing olfactory training twice a day was too often?”, *X*^*2*^ (1, *N* = 99) = 32.07, *p* <.001, but not to the questions “Did you stop performing olfactory training on your own accord, because you felt that your sense of smell did NOT improve?”, *X*^*2*^ (1, *N* = 99) = .018, *p* = .89, and “Did you stop performing olfactory training on your own accord because you felt that your sense of smell DID improve?”, *X*^*2*^ (1, *N* = 99) < .001, *p* = 1, Figure 4. Critically, in response to the question “How often did you forget to perform smell training”, significantly fewer individuals in the NI group reported forgetting to train, *X*^*2*^ (4, *N* = 99) = 26.51, *p* < .001. In fact, as stated above, a full 98% reported that they never missed a training session when training with the nasal inserts. This indicates that participants in the nasal insert group were more consistent with their olfactory training.

**Figure 4.**
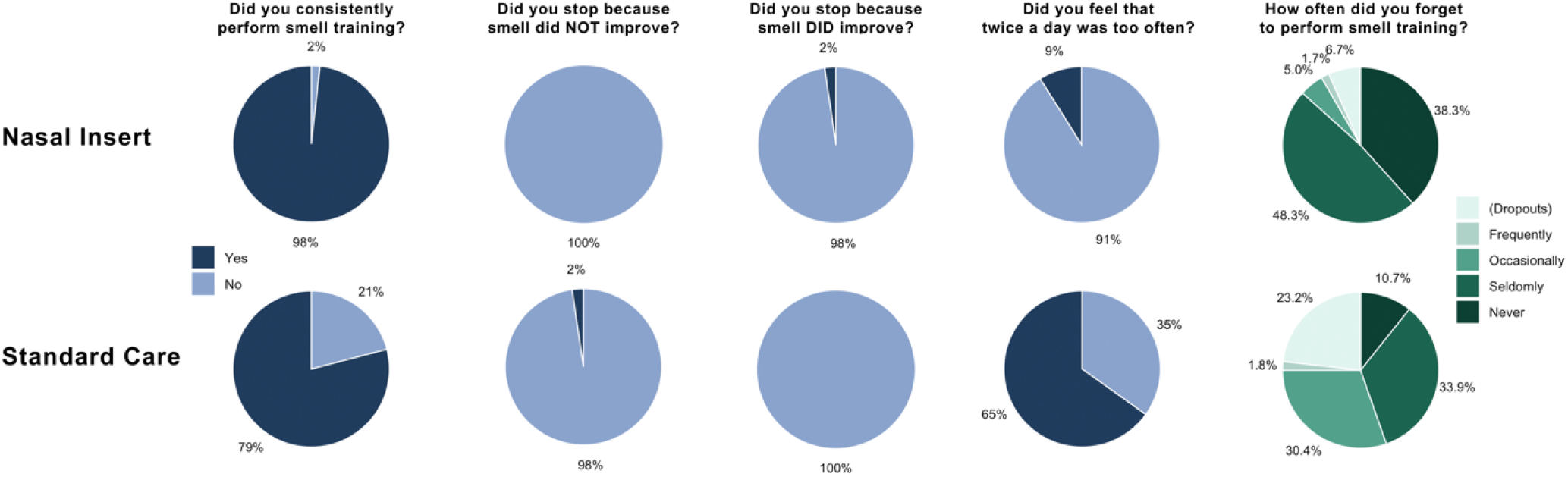
Frequency of answers to the adherence questionnaire separated by olfactory training group.

## DISCUSSION

In this study, we aimed to investigate how a modified version of olfactory training using scented nasal inserts compares to standard olfactory training in treatment effect and compliance rate. Using a technique where odors are administered using scented intranasal plugs, we could demonstrate a significantly higher adherence to treatment protocol with more consistent training and fewer forgotten training session while maintaining treatment effects, compared to standard care treatment. Critically, training with nasal inserts reduced dropout rates to a mere 6.7% compared to 23.2% in the standard care group.

Individuals in the nasal insert training group demonstrated an overall greater adherence to treatment and higher satisfaction. Of those completing the study, nearly 98% of the nasal insert training group reported that they consistently performed their training compared to 79% in the standard care group and a full 86.6% reported that they seldom or never forgot a session, compared to 64.3% in the standard care group. The exact reasons why the standard care group reported lower adherence to treatment protocol is difficult to determine but 35% of the group reported that training twice a day was too much compared to 9% in the nasal insert group. It can be speculated that the increased adherence in the nasal insert group might be attributed to the enhanced mobility during training that the inserts allowed them. Alternatively, the difference in the number of odors used for training (6 SC vs 10 NI) might have contributed given that a recent study suggests that increasing from 4 to 6 odors increase adherence to treatment protocol (Genetzaki et al., 2024). A more simplistic explanation is that the mere act of being provided with some sort of medical system increases the perceived saliency of the training. The exact mediating mechanism notwithstanding, in all medical treatment, adherence to treatment protocol and persistence in maintain prescribed treatment is a key factor in treatment outcomes on both the individual and group level. Past studies have demonstrated that only half of all medical patients adhere to the medical advice they receive from their attending doctor (M R DiMatteo, 1994) and close to 50% of patients do not take their prescribed medication as frequent as instructed, or at all (Epstein & Cluss, 1982). Medical treatment requiring even more time and effort from the patient, such as at home physical therapy regimens, have a non-adherence rate as high as 70% (Sluijs et al., 1993). Adherence to olfactory training in the general clinical population is similarly suboptimal where at least 15% of patients do not even start their recommended training regimen and among those who start, only 33% self-report consistent training (Haas et al., 2024). Considering both the lower dropout rates and higher adherence to treatment for those following through with the treatment in the nasal insert group, it can be speculated that it is a potential clinically advantageous method to increase overall effectiveness of odor training in a clinical population.

There were significantly fewer dropouts in the nasal insert group (6.7% vs 23.2%). While this is potentially positive for treatment outcome, we do not know exactly why someone discontinues a study. Individuals may drop-out from a study because they do not experience any effect or because they experience that they are cured and do not need further treatment. It is worth highlighting that we used a validated adherence to treatment scale that was developed specifically for odor training (Haas et al., 2024), but the two questions pertaining to why participants choose to discontinue are less informative in the context of this study because only individuals who attended the final visit answered them. Nonetheless, past studies suggest that individuals without any self-perceived improvements are more likely to discontinue olfactory training (Haas et al., 2024).

A limitation of the present study is that we could not control that the participants in the nasal insert group focused on the odor object, represented by an image on the individual package, as instructed, rather than some other task at hand. Because active mentalization of the odor object in question has been demonstrated to exchange olfactory training outcome (Pieniak et al., 2022), this might potentially mask a potential difference in treatment outcome between groups. However, this would mean that the nasal inserts improvement could be potentially larger and therefore do not unduly bias our results. Moreover, we only included participants with post-viral or idiopathic hyposmia, meaning that we cannot generalize our results to other patient groups with olfactory dysfunction. Finally, the current recommended duration of olfactory training in the Swedish healthcare system is three months and it is known that longer periods of training yield better outcomes (Konstantinidis et al., 2016). It could therefore be speculated that olfactory function could have increased had the length of training been extended.

In conclusion, we demonstrate that olfactory training with scented nasal inserts leads to a significantly higher adherence to treatment protocol with more consistent training and fewer forgotten training sessions while maintaining treatment effects, when compared to standard care. The combination of significantly lower dropout rates and higher adherence, while maintaining treatment outcomes, makes nasal inserts an interesting method to increase the effectiveness of olfactory training in standard clinical populations.

## Data Availability

Upon journal acceptance, all raw data and analyses scripts will be available at the Open Science Framework (OSF).

https://osf.io/g6e53/?view_only=60e7df87bd8647a389630dfa55b6b527

## ACKNOWLEDGEMENTS

We thank Pia Sandell for helping with testing.

## AUTHORSHIP CONTRIBUTION

JNL, ALW, and PSJ contributed to conception and design of the study. ALW together with MS and JM refined the study protocol and collected the data. ALW performed the statistical analysis and wrote the first draft of the manuscript. JNL, SH, ET, and PSJ wrote sections of the manuscript. All authors edited versions of the manuscript, read, and approved the submitted version.

